# Donor genetic variants in interleukin-6 and interleukin-6 receptor associate with biopsy-proven rejection following kidney transplantation

**DOI:** 10.1101/2021.04.17.21255669

**Authors:** Felix Poppelaars, Mariana Gaya da Costa, Siawosh K. Eskandari, Jeffrey Damman, Marc A. Seelen

**Affiliations:** Department of Internal Medicine, Division of Nephrology, University Medical Center Groningen, University of Groningen, Groningen, The Netherlands; Department of Pathology, Erasmus Medical Center, University Medical Center, Rotterdam, The Netherlands

**Keywords:** Cytokines, Kidney, Transplantation, Nephrology

## Abstract

Rejection after kidney transplantation remains an important cause of allograft failure that markedly impacts morbidity. Cytokines are a major player in rejection, and we, therefore, explored the impact of interleukin-6 (*IL6*) and IL-6 receptor (*IL6R*) gene polymorphisms on the occurrence of rejection after renal transplantation. We performed an observational cohort study analyzing both donor and recipient DNA in 1,271 renal transplant-pairs from the University Medical Center Groningen in The Netherlands and associated single nucleotide polymorphisms (SNPs) with biopsy-proven rejection after kidney transplantation. The C-allele of the *IL6R* SNP (Asp358Ala; rs2228145 A>C, formerly rs8192284) in donor kidneys conferred a reduced risk of rejection following renal transplantation (HR 0.78 per C-allele; 95%-CI 0.67–0.90; *P*=0.001). On the other hand, the C-allele of the *IL6* SNP (at position-174 in the promoter; rs1800795 G>C) in donor kidneys was associated with an increased risk of rejection for male organ donors (HR per C-allele 1.31; 95%-CI 1.08–1.58; *P*=0.0006), but not female organ donors (*P*=0.33). In contrast, neither the *IL6* nor *IL6R* SNP in the recipient showed an association with renal transplant rejection. In conclusion, donor *IL6* and *IL6R* genotypes but not recipient genotypes represent an independent prognostic marker for biopsy-proven renal allograft rejection.

## Introduction

Since the first successful kidney transplant in 1954, kidney transplantation has become the treatment of choice for patients with end-stage kidney disease (ESKD).^1^ Beyond the surgical advances in performing kidney transplantation, it was the addition of immunosuppressive drugs post-transplant that improved the survival of kidney allografts from months to years. Transplant recipients, however, invariably become sensitized to their transplant, leading to poor long-term survival of kidney allografts.^2,3^ While conventional immunosuppressants are effective at abolishing acute immune responses, they only partially address allosensitization—thus allowing humoral immunity to form and precipitate long-term allograft failure. To prevent rejection of kidney transplants it is thus necessary to develop novel therapies that can impede the formation of allosensitization.^4^

Through the scientific efforts of the past fifty years, it has been established that acute and chronic allograft rejection are triggered by a complex and dynamic set of alloimmune responses. Immunomodulatory proteins known as cytokines are particularly important in regulating both the pro- and anti-inflammatory arms of immunity. Among all cytokine superfamilies, interleukin 6 (IL-6) and its family members arguably show the greatest degree of cytokine functional pleiotropy (multiple biological actions) and cytokine redundancy (shared biological actions), with physiological and pathological roles ranging from B and T cell differentiation and acute-phase immunity, to hematopoiesis and oncogenesis, tissue and bone remodeling and regeneration, and even early development of neuronal and cardiovascular systems.^5–8^ As such, aberrant IL-6 activity is implicated in diseases including systemic inflammatory response syndrome, chronic immune disorders such as crescentic glomerulonephritis, transplant rejection, and graft-versus-host disease, rheumatic diseases including rheumatoid arthritis and juvenile idiopathic arthritis, and lymphoproliferative conditions like Castleman’s disease.^4,8,9^

Emerging data has identified a key role for IL-6 in rejection after renal transplantation.^4^ In order to appreciate the role of IL-6 signalling in renal allograft rejection, it is of value to contextualize the signalling cascade first as IL-6 can signal through three separate pathways and command varying effects depending on the context and the pathway at play.^5,6,10^ The first mode of signaling is known as classic signaling, in which IL-6 binds to membrane-bound IL-6 receptor (mIL-6R) followed by complex formation with membrane-bound glycoprotein 130 (gp130) to trigger downstream signal transduction and gene expression. Trans signaling instead occurs when the IL-6R is cleaved from the cell membrane,^10^ and the released soluble IL-6R (sIL-6R) captures IL-6, forms a complex with membrane-bound gp130, and phosphorylates intracellular signal transducers. Since membrane-bound gp130 is ubiquitously expressed but mIL-6R is not, trans signaling can facilitate IL-6 responsivity in cells that would not normally respond to IL-6 through IL-6/sIL-6R complexes, extending the physiological and pathological functions of IL-6.^7^ Importantly, a soluble form of gp130 naturally exists in circulation with a similar affinity for the IL-6/sIL-6R complex as membrane-bound gp130 and can act as a specific inhibitor of the trans signaling pathway.^11^ The third and final form of IL-6 signaling is trans presentation where mIL-6R on one cell captures IL-6 and binds to membrane-bound gp130 on another cell. In all these modes of signaling, phosphorylation of secondary messengers of particularly the Janus kinase (JAK) and signal transducer and activator of transcription (STAT) families are critical for enacting IL-6-dependent effects.^9,14^

Here, we used human genetics to investigate whether blockade of IL-6/IL-6R interaction might confer therapeutic benefit in renal transplantation. The IL-6-related single nucleotide polymorphisms (SNPs) we studied here are the A>C polymorphism in *IL6R* (rs2228145) and the G>C polymorphism in *IL6* (rs1800795). The *IL6R* SNP causes a missense polymorphism leading to increased concentrations of sIL-6R in the circulation (inhibiting the trans signaling pathway), while the *IL6* SNP causes elevated levels of IL-6 expression due to its location in a promoter region (promoting IL-6 signaling via all three pathways).^14^ Other studies have previously assessed the role of this *IL6* variants in renal transplantation, finding associations between *IL6* variants and transplant outcome.^12,13^ These studies, however, are mostly retrospective, limited in their statistical methods, underpowered and therefore inconclusive due to the limited number of patient samples included in the analyses. In the present study, we provide a decisive answer on the impact of these SNPs in kidney transplantation.

## Results

### Determinants of biopsy-proven rejection following kidney transplantation

A total of 1,271 kidney transplant donor-recipient pairs were included in this study with the donor and recipient characteristics listed in Table 1. The mean follow-up after transplantation was 5.2 years ± 5.0 with a maximum follow-up period of 16.7 years. During follow-up, 33.8% of the recipients developed biopsy-proven rejection. In the first year, 390 recipients developed acute biopsy-proven rejection, whereas 40 recipients presented with biopsy-proven rejection thereafter. Of all the assessed characteristics, the following were significantly associated with biopsy-proven rejection (Table 1); recipient age, recipient sex, warm ischemia time, the total number of HLA mismatches, and the occurrence of delayed graft function (DGF).

**Table 1:** Baseline characteristics of the donors and recipients.

|  | All Patients<br>(n = 1271) | No rejection<br>(n = 839) | Rejection<br>(n = 432) | P-value* | HR | P-value# |
| --- | --- | --- | --- | --- | --- | --- |
| Donor |  |  |  |  |  |  |
| Age, years | 44.4 ± 14.4 | 44.8 ± 14.6 | 43.6 ± 14.1 | 0.15 |  | 0.17 |
| Male sex, n (%) | 645 (50.7) | 423 (50.4) | 222 (51.4) | 0.74 |  | 0.77 |
| Blood group |  |  |  |  |  |  |
| Type O, n (%) | 642 (50.5) | 421 (50.4) | 221 (51.2) | 0.16 |  | 0.20 |
| Type A, n (%) | 502 (39.5) | 343 (41.0) | 159 (36.8) |  |  |  |
| Type B, n (%) | 97 (7.6) | 55 (6.6) | 42 (9.7) |  |  |  |
| Type AB, n (%) | 27 (2.1) | 17 (2.0) | 10 (2.3) |  |  |  |
| Donor type |  |  |  |  |  |  |
| Living, n (%) | 282 (22.2) | 190 (22.6) | 92 (21.3) | 0.40 |  | 0.41 |
| Brain death, n (%) | 787 (61.9) | 509 (60.7) | 278 (64.4) |  |  |  |
| Circulatory death, n (%) | 202 (15.9) | 140 (16.7) | 62 (14.4) |  |  |  |
| Cause of death |  |  |  |  |  |  |
| Trauma, n (%) | 305 (30.8) | 353 (42.1) | 196 (45.4) | 0.57 |  | 0.57 |
| CVA, n (%) | 549 (55.5) | 207 (24.7) | 98 (22.7) |  |  |  |
| Other, n (%) | 135 (13.7) | 279 (33.2) | 138 (31.9) |  |  |  |
| Recipient |  |  |  |  |  |  |
| Age, years | 47.9 ± 13.5 | 49.6 ± 13.4 | 44.5 ± 12.9 | <0.0001 | 0.98 | <0.0001 |
| Male sex, n (%) | 739 (58.1) | 469 (55.9) | 270 (62.5) | 0.02 | 0.80 | 0.03 |
| Primary kidney disease |  |  |  |  |  |  |
| Glomerulonephritis, n (%) | 340 (26.8) | 214 (25.5) | 126 (29.2) | 0.28 |  | 0.25 |
| Polycystic disease, n (%) | 208 (16.4) | 139 (16.6) | 69 (16.0) | 0.58 |  | 0.51 |
| Vascular disease, n (%) | 145 (9.9) | 102 (12.2) | 43 (10.0) | 0.19 |  | 0.18 |
| Pyelonephritis, n (%) | 148 (11.6) | 97 (11.6) | 51 (11.8) | 1.00 |  | 0.93 |
| Diabetes, n (%) | 51 (4.0) | 31 (3.7) | 20 (4.6) | 0.55 |  | 0.44 |
| Chronic, n (%) | 168 (13.2) | 107 (12.8) | 61 (14.1) | 0.66 |  | 0.71 |
| Other, n (%) | 231 (18.2) | 149 (17.8) | 62 (14.4) | 0.50 |  | 0.60 |
| Blood group |  |  |  |  |  |  |
| Type A, n (%) | 536 (42.2) | 372 (44.3) | 195 (45.1) | 0.41 |  | 0.62 |
| Type B, n (%) | 113 (8.9) | 364 (43.4) | 172 (39.8) |  |  |  |
| Type O, n (%) | 567 (44.6) | 71 (8.5) | 42 (9.7) |  |  |  |
| Type AB, n (%) | 55 (4.3) | 32 (3.8) | 23 (5.3) |  |  |  |
| Dialysis vintage, weeks | 189 ± 136 | 184 ± 139 | 198 ± 129 | 0.09 |  | 0.06 |
| Highest PRA, in % | 10.1 ± 23.6 | 10.1 ± 23.4 | 10.3 ± 23.8 | 0.86 |  | 0.81 |
| Immunosuppression |  |  |  |  |  |  |
| Azathioprine, n (%) | 72 (5.7) | 44 (5.2) | 28 (6.5) | 0.37 |  | 0.22 |
| Corticosteroids, n (%) | 1201 (94.5) | 787 (93.8) | 414 (95.8) | 0.13 |  | 0.16 |
| Cyclosporin, n (%) | 1085 (85.4) | 717 (85.5) | 368 (85.2) | 0.90 |  | 0.82 |
| Mycophenolic acid, n (%) | 907 (71.4) | 611 (72.8) | 296 (68.5) | 0.11 |  | 0.10 |
| Sirolimus, n (%) | 38 (3.0) | 23 (2.7) | 15 (3.5) | 0.47 |  | 0.43 |
| Tacromilus, n (%) | 97 (7.6) | 61 (7.3) | 36 (8.3) | 0.50 |  | 0.38 |
| Transplantation |  |  |  |  |  |  |
| CIT, in hours | 16.7 ± 9.4 | 16.4 ± 9.3 | 17.3 ± 9.8 | 0.09 |  | 0.15 |
| WIT, in minutes | 39.0 ± 11.3 | 39.6 ± 12.0 | 37.9 ± 9.9 | 0.005 | 0.99 | 0.009 |
| Total HLA mismatches | 2 [1 – 3] | 2 [0 – 3] | 2 [1 – 3] | <0.0001 | 1.20 | <0.0001 |
| DGF, n (%) | 415 (32.7) | 256 (30.5) | 159 (36.8) | 0.02 | 1.30 | 0.008 |

### A common genetic variant in interleukin-6 receptor protects against renal allograft rejection

To identify relevant genetic biomarkers in the context of rejection, we explored whether the Asp358Ala variant in *IL6R* (rs2228145 A>C, previously rs8192284) associated with biopsy-proven rejection. The observed genotype frequencies in recipients (n=1270; AA, 34.3%; AC, 51.4%; CC, 14.2%) and donors (n=1269; AA, 35.6%; AC, 50.6%; CC, 13.6%) were comparable (*P* = 0.75). Yet, the frequencies of the *IL6R* polymorphism in both recipients and donors were significantly higher than reported by the 1000 genomes project (P<0.0001), but not when compared to their European cohort (*P* = 0.09).^15^ Kaplan– Meier survival analyses showed that the *IL6R* polymorphism in the donor was significantly associated with a reduced risk for acute rejection during follow-up (Fig. 1A; log-rank *P* = 0.003). After the complete follow-up, the incidence of biopsy-proven rejection was 38.9% in the reference AA genotype group, 32.8% in the heterozygous AC genotype group, and 24.9% in the homozygous CC genotype group, respectively. In the recipients, Kaplan–Meier survival analyses revealed no associations between the *IL6R* SNP and rejection during follow-up (Fig 1B; log-rank *P* = 0.70). Subgroup analysis for recipient sex or donor type did not change these results. In univariate analysis, the *IL6R* SNP in the donor was associated with a hazard ratio of 0.78 per C-allele (95%-CI: 0.67 – 0.90; *P* = 0.001) for biopsy-proven rejection. A multivariable analysis was performed to adjust for potential confounders (Table 2), including donor characteristics (model 2), recipient characteristics (model 3), and transplant variables (model 4). In Cox regression analysis, the *IL6R* SNP in the donor remained significantly associated with biopsy-proven rejection independent of potential confounders. Finally, we performed a multivariable analysis with a stepwise forward selection procedure using all variables associated with rejection in univariable analysis (Table 3). In the final model, the *IL6R* SNP in the donor, recipient age and sex, the total number of HLA mismatches, and DGF were included. After adjustment, the *IL6R* SNP in the donor was associated with rejection with a hazard ratio of 0.73 per C-allele (95% CI: 0.62 – 0.86, *P*<0.001). Hence, altogether these results demonstrate that a common functional variant in the *IL6R* gene in the donor associates with a lower incidence of biopsy-proven acute rejection after kidney transplantation.

**Table 2.** Associations of interleukin-6 and interleukin-6 receptor polymorphisms in the donor with rejection following kidney transplantation.

|  | *IL6R* SNP (rs2228145-C*) in the donor |  |  | *IL6* SNP (rs1800795-C) in male donors |  |  |
| --- | --- | --- | --- | --- | --- | --- |
|  | Hazard ratio<br>(per allele) | 95% CI | P-value | Hazard ratio<br>(per allele) | 95% CI | P-value |
| **Model 1** | 0.78 | 0.67 – 0.90 | 0.001 | 1.31 | 1.08 – 1.58 | 0.006 |
| **Model 2** | 0.77 | 0.67 – 0.89 | 0.001 | 1.31<sup>#</sup> | 1.08 – 1.59 | 0.005 |
| **Model 3** | 0.75 | 0.65 – 0.88 | <0.001 | 1.29 | 1.06 – 1.56 | 0.01 |
| **Model 4** | 0.74 | 0.63 – 0.87 | <0.001 | 1.30 | 1.06 – 1.61 | 0.01 |

Data are presented as hazard ratio with 95% confidence interval (CI) and P-value.
**Model 1:** crude model
**Model 2:** adjusted for model 1 plus donor characteristic's: donor age, donor sex, donor blood type, and donor origin.
**Model 3:** adjusted for model 1 plus recipient characteristic's: recipient age, recipient sex, recipient blood type and dialysis vintage.
**Model 4:** adjusted for model 1 plus transplant characteristic's: cold and warm ischemia time, the total HLA-mismatches, and the occurrence of delayed graft function (DGF).
\* Previously rs8192284
<sup>#</sup>For the *IL6* SNP (rs1800795-C), model 2 could not be adjusted for donor sex since this was a subgroup analysis for male donors.

**Table 3.** Competitive analysis of the associations of clinical factors with rejection following kidney transplantation.

| Variables not in the equation |  | Variables in the equation |  |  |
| --- | --- | --- | --- | --- |
| Variables | P-value | Variables | P-value | Hazard Ratio |
| Dialysis vintage | 0.97 | rs2228145-C In the donor | <0.001 | 0.73 (0.62 – 0.86) |
| Warm ischemia time | 0.05 | Total HLA-mismatches | <0.001 | 1.19 (1.10 – 1.29) |
|  |  | Recipient age | <0.001 | 0.97 (0.97 – 0.98) |
|  |  | DGF | 0.023 | 1.28 (1.04 – 1.59) |
|  |  | Recipient sex | 0.023 | 0.78 (0.63 – 0.97) |

Multivariable cox regression was performed with a stepwise forward selection. Only variables that with a P-value below 0.10 in the univariate analysis were included. Data are presented as hazard ratio with 95% confidence interval (CI) and P-value. In the final, the *IL6R* SNP (rs2228145-C, previously rs8192284) In the donor, total number of HLA-mismatches, recipient age, the occurrence of delayed graft function (DGF) and recipient sex were included, whereas dialysis vintage and warm ischemia time were not.

**Figure 1.**
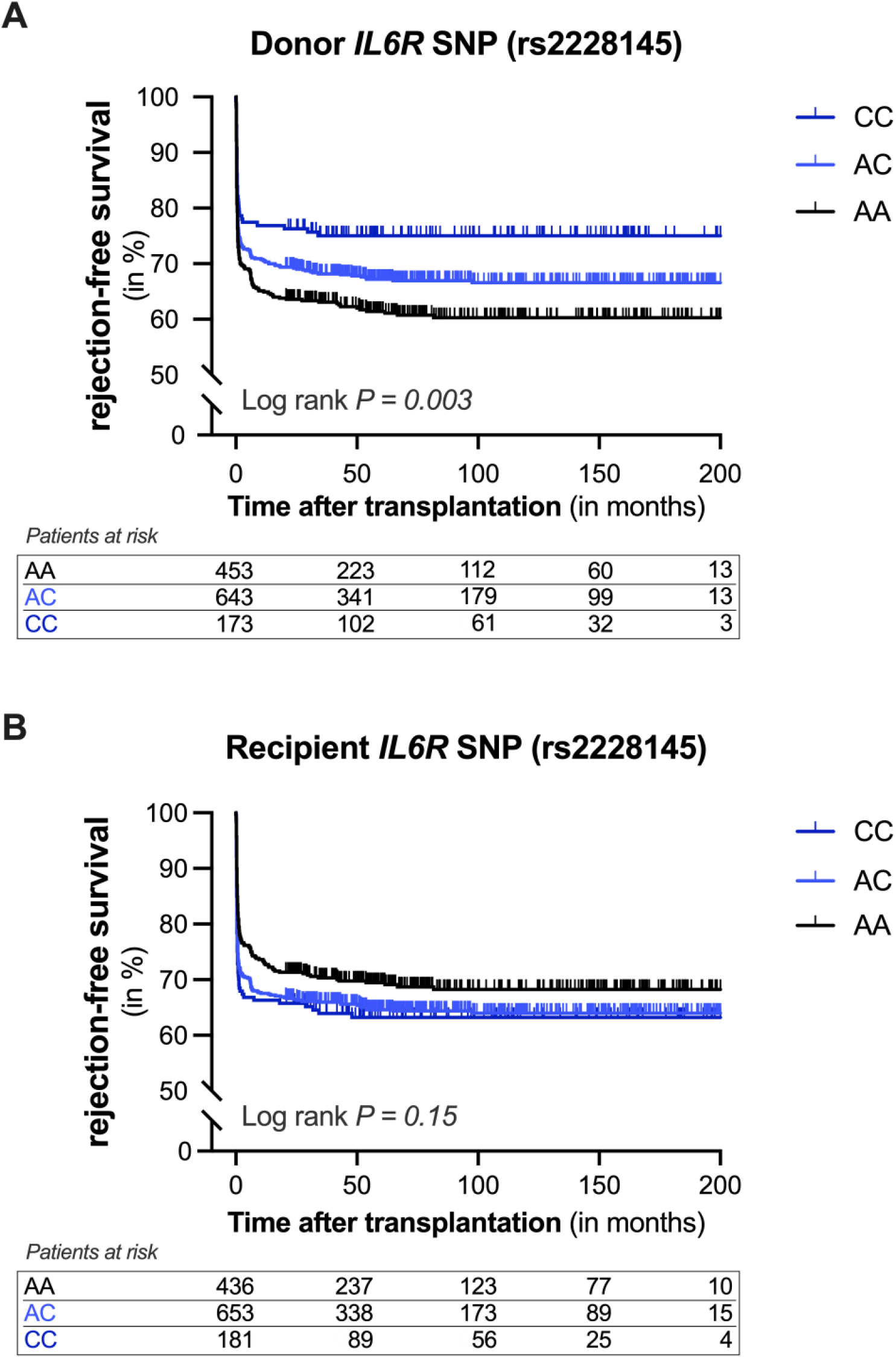
**Kaplan-Meier curves for rejection-free survival of kidney allografts according to the presence of the interleukin-6 receptor variant in the donor or recipient**. Cumulative rejection-free survival of renal allografts according to the presence of the Asp358Ala variant in IL-6 receptor (*IL6R*, rs2228145 A>C, previously rs8192284) in (A) the donor, and (B) the recipient. Log-rank test was used to compare the incidence of biopsy-proven rejection between the groups.

### A common genetic variant in interleukin-6 is a risk factor for renal allograft rejection

Next, we explored whether the *IL6* SNP at position-174 in the promoter (rs1800795 G>C) affects the incidence of biopsy-proven rejection. The observed genotype frequencies in the recipients (n=1268: GG, 37.8%; GC, 47.0%; CC, 14.9%) and donors (n=1260: GG, 33.1%; AC, 49.3%; CC, 16.8%) differed significantly (*P* = 0.049). More specifically, the C-allele of the *IL6* SNP seemed to be more prevalent in the kidney donors. The frequencies of the *IL6* SNP in both the recipients and donors did not significantly differ from those reported by the European cohort of the 1000 genomes project (*P* = 0.09 and *P* = 0.19, respectively).^15^ Kaplan–Meier analysis demonstrated that neither the *IL6* SNP in the donor affected the risk of rejection (Fig. 2A, *P* = 0.58) nor the *IL6* SNP in the recipient (Fig. 2B, *P* = 0.70). In additional analysis, the *IL6* SNP in the recipient was not tied to the risk of biopsy-proven rejection within the first year either (*P* = 0.19).

**Figure 2.**
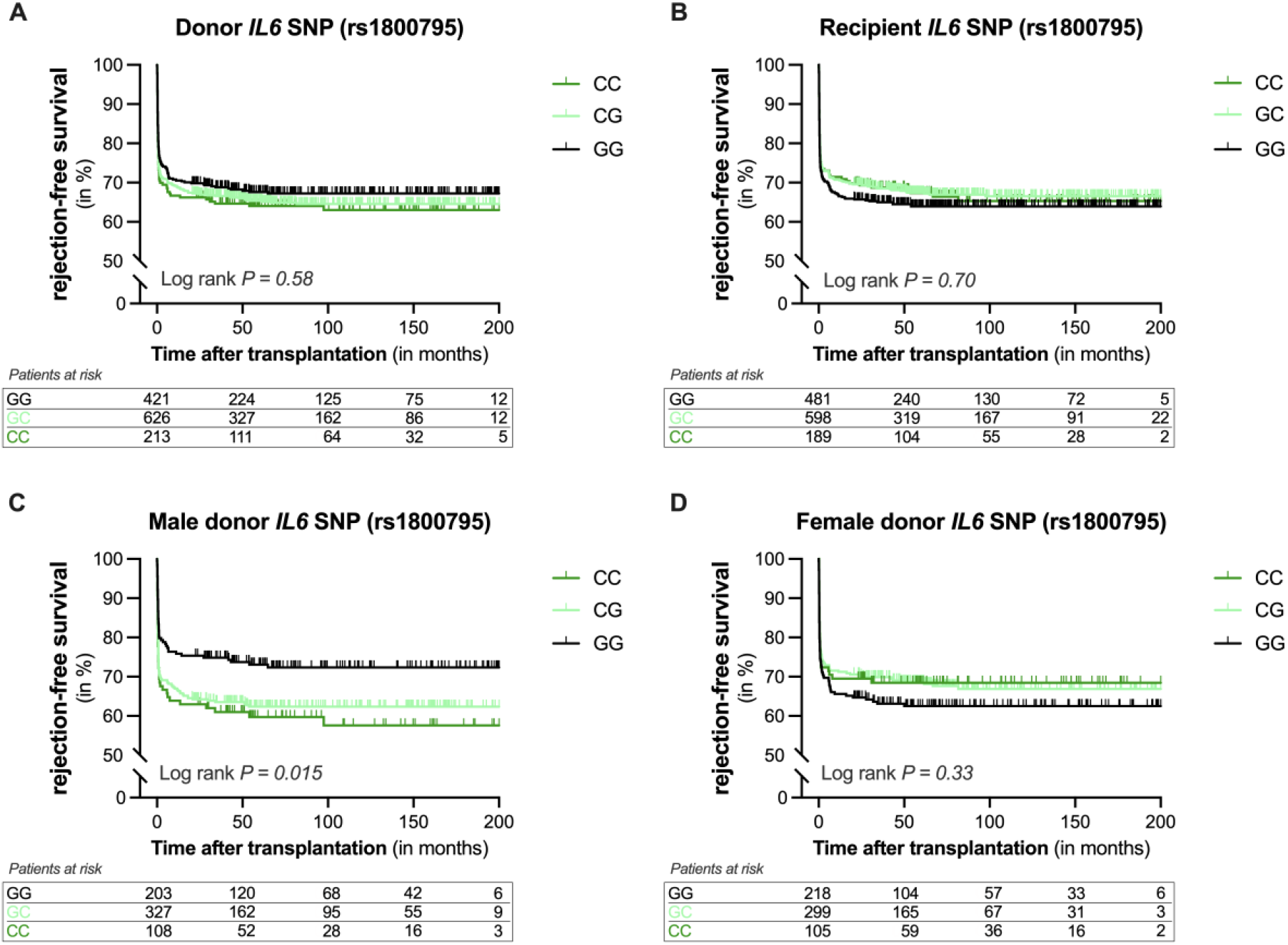
**Kaplan-Meier curves for rejection-free survival of kidney allografts according to the presence of the interleukin-6 genetic variant in the donor or recipient**. Cumulative rejection-free survival of renal allografts according to the presence of the interleukin-6 polymorphism (*IL6*) at position-174 in the promoter (rs1800795 G>C) in (A) the donor and (B) the recipient. A subgroup analysis was performed for donor sex and cumulative rejection-free survival was shown according to the presence of the *IL6* SNP in (C) male donors and (D) female donors. Log-rank test was used to compare the incidence of biopsy-proven rejection between the groups.

We next performed a subgroup analysis for sex, since sex is known to impact immunity, renal disease as well as transplantation outcome.^16,17^ Kaplan-Meier curves revealed a significant association between the *IL6* SNP and rejection in male donors (Fig. 2C, *P* = 0.015). After the complete follow-up, the incidence of biopsy-proven rejection was 27.1% in the reference GG genotype group, 37.3% in the heterozygous GC genotype group, and 40.7% in the homozygous CC genotype group, respectively. No association was seen between the *IL6* SNP and rejection in female donors (Fig. 2D, *P* = 0.33). Univariate analysis revealed that the *IL6* SNP in male donors was associated with a hazard ratio of 1.31 per C-allele (95%-CI: 1.08 – 1.58; *P* = 0.006) for biopsy-proven rejection. Multivariable analysis was performed to adjust for potential confounders (Table 2), including donor characteristics (model 2), recipient characteristics (model 3), and transplant variables (model 4). In Cox regression analysis, the *IL6* SNP in male donors remained significantly associated with biopsy-proven rejection independent of potential confounders. Thus, it appears that a functional variant in *IL6* in male donors is linked to a higher incidence of biopsy-proven rejection after kidney transplantation.

### Prediction of biopsy-proven rejection

The combined presence of the *IL6* and *IL6R* SNP was common (Fig. 3A – B), as well as the presence of the same SNP in the donor and recipient of a transplant pair (Fig. 3C – D). We, therefore, speculated that assessing the combined presence of multiple polymorphisms, a genetic risk score, in the donor-recipient pairs could yield more information than examining the polymorphisms individually. To explore the combination of *IL6* and *IL6R* SNPs as predictors of biopsy-proven rejection, we created a genetic risk score of the two variants in both the donors and recipients. Weight was added to each SNP according to their hazard ratio, creating a negative score for protective polymorphisms and a positive score for hazardous ones. Overall, a genetic risk score above zero indicates the presence of more hazardous SNPs in a donor-recipient pair and genetic risk scores below zero indicate the presence of more protective SNPs in a donor-recipient pair. To assess the clinical applications of the IL-6/IL-6R genetic risk score, we studied the predictive value of this genetic profiling in more detail. The genetic risk score was significantly associated with biopsy-proven rejection in both the crude model (HR, 1.24; 95%-CI, 1.12 – 1.36; *P*<0.001 per SD increase) and after adjustment for donor characteristics, recipient characteristics, and transplant variables (Table 4). Furthermore, the hazard ratio of the IL-6/IL-6R genetic risk score was consistent in subgroups analyses and remained significant (Figure 4), except in living donors after stratification for donor type (*P* = 0.22). The confidence intervals of all subgroups showed substantial overlap with the overall hazard ratio at the top, demonstrating the consistency of the findings across subgroups. Next, we tested if the IL-6/IL-6R genetic risk score was a better predictor for rejection than the IL-6R SNP in the donor by multivariable regression with a stepwise forward selection was performed (Table 5). In the final model, the IL-6/IL-6R genetic risk score was included whereas the *IL6R* SNP in the donor was not. After adjustment, the genetic risk score in the donor was associated with biopsy-proven rejection with a hazard ratio of 1.25 per SD increase (Figure 5, 95% CI: 1.13 – 1.39; *P*<0.001).

**Table 4.** Associations of interleukin-6/interleukin-6 receptor genetic risk score with rejection following kidney transplantation.

|  | IL-6/IL-6R genetic risk score |  |  |
| --- | --- | --- | --- |
|  | Hazard ratio<br>(per SD) | 95% CI | P-value |
| **Model 1** | 1.24 | 1.12 – 1.36 | <0.001 |
| **Model 2** | 1.23 | 1.12 – 1.36 | <0.001 |
| **Model 3** | 1.26 | 1.14 – 1.39 | <0.001 |
| **Model 4** | 1.25 | 1.13 – 1.39 | <0.001 |

Data are presented as hazard ratio with 95% confidence interval (CI) and P-value.
**Model 1:** crude model
**Model 2:** adjusted for model 1 plus donor characteristic's: donor age, donor sex, donor blood type, and donor origin.
**Model 3:** adjusted for model 1 plus recipient characteristic's: recipient age, recipient sex, recipient blood type and dialysis vintage.
**Model 4:** adjusted for model 1 plus transplant characteristic's: cold and warm ischemia time, the total HLA-mismatches, and the occurrence of delayed graft function (DGF).

**Table 5.** Competitive analysis of the associations of clinical factors with rejection following kidney transplantation.

| Variables not in the equation |  | Variables in the equation |  |  |
| --- | --- | --- | --- | --- |
| Variables | P-value | Variables | P-value | Hazard Ratio |
| rs2228145-C* In the donor | 0.68 | IL-6/IL-6R genetic risk score | <0.001 | 1.27 (1.14 – 1.42) |
| Dialysis vintage | 0.97 | Total HLA-mismatches | <0.001 | 1.19 (1.10 – 1.29) |
|  |  | Recipient age | <0.001 | 0.97 (0.97 – 0.98) |
|  |  | DGF | 0.008 | 1.34 (1.08 – 1.67) |
|  |  | Recipient sex | 0.031 | 0.79 (0.63 – 0.98) |
|  |  | Warm ischemia time | 0.049 | 0.99 (0.98 – 1.00) |

Multivariable cox regression was performed with a stepwise forward selection. Only variables that with a P-value below 0.10 in the univariate analysis were included. Data are presented as hazard ratio with 95% confidence interval (CI) and P-value. The IL-6/IL-6R genetic risk score, total number of HLA-mismatches, recipient age, occurrence of delayed graft function (DGF), recipient sex and warm ischemia time were included in the final model.
\* Previously rs8192284

**Figure 3.**
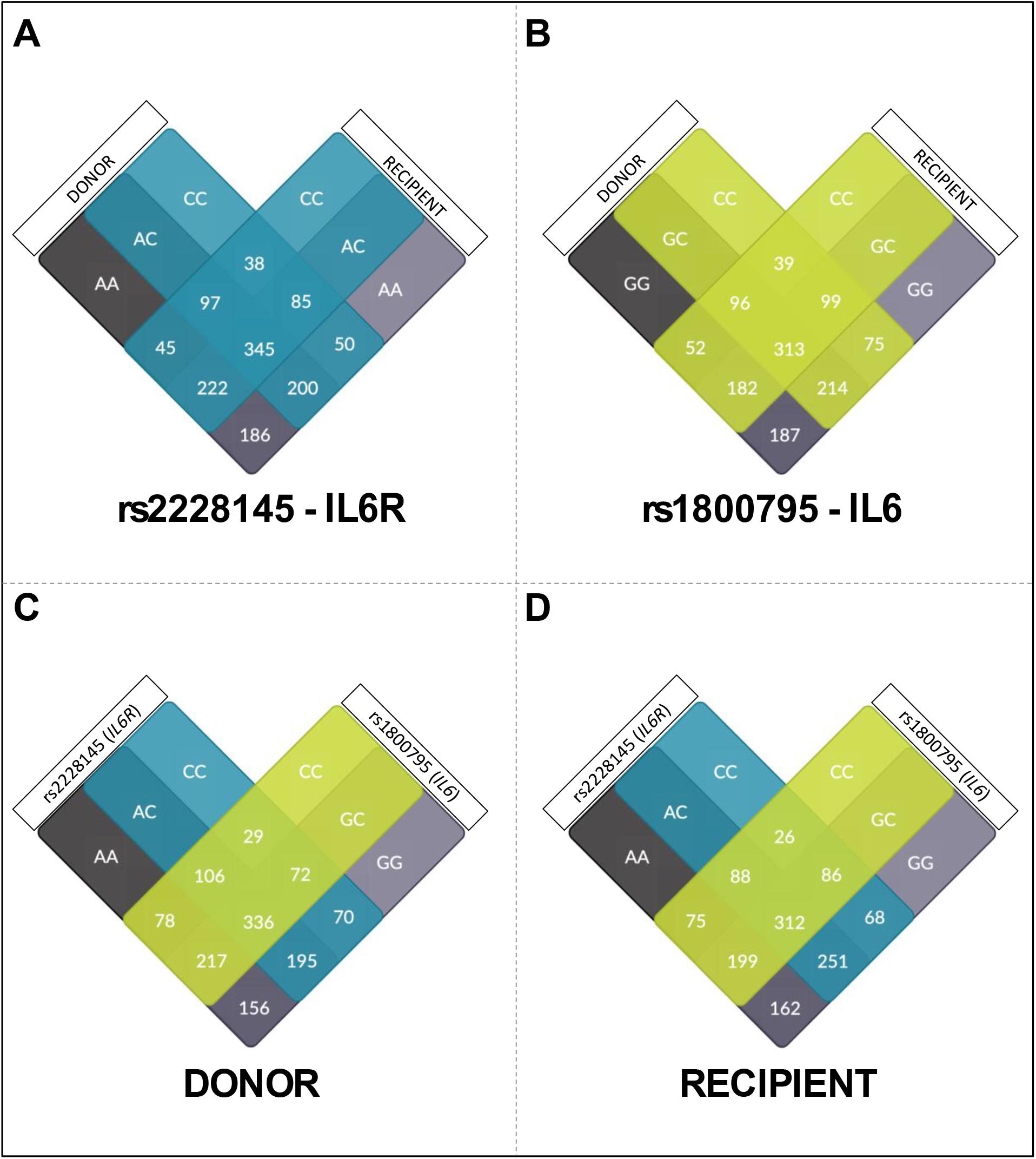
Venn diagram of the genetic status of the two polymorphisms in the donor, recipient and pairs. 1271 donor-recipient renal transplant pairs were analyzed for the presence of a polymorphism in interleukin-6 (*IL6*) at position-174 in the promoter (rs1800795 G>C) and Asp358Ala variant in IL-6 receptor (*IL6R*, rs2228145 A>C, previously rs8192284). The Venn diagram depicts the number of renal transplant pairs based on their (A) *IL6R* genotype or (B) *IL6* genotype. In addition, the combined genotype of *IL6* and *IL6R* are depicted in (C) the donor and (D) the recipient.

**Figure 4.**
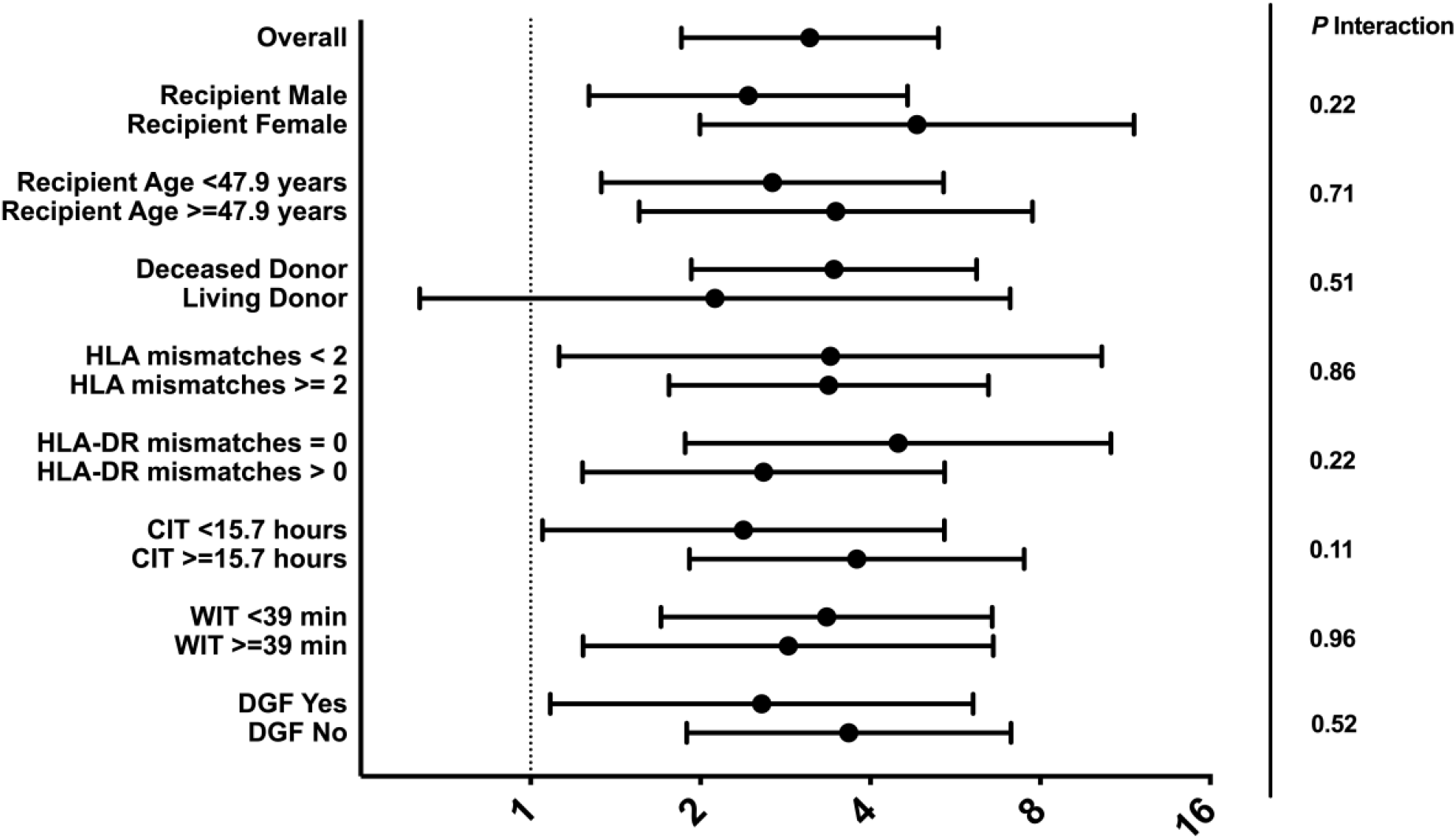
Hazard ratios for the interleukin-6/interleukin-6 receptor genetic risk score among subgroups. Forest plot of sub-analyses of the IL-6/IL-6R genetic risk score, demonstrating that the hazard ratios for biopsy-proven rejection were consistent in different subgroups. The only exception was the donor origins of the kidney allografts. The association between the IL-6/IL-6R genetic risk score and rejection was not seen in kidney transplants from living donors. No significant interaction was seen between the IL-6/IL-6R genetic risk score and the different clinical variables of the subgroups.

**Figure 5.**
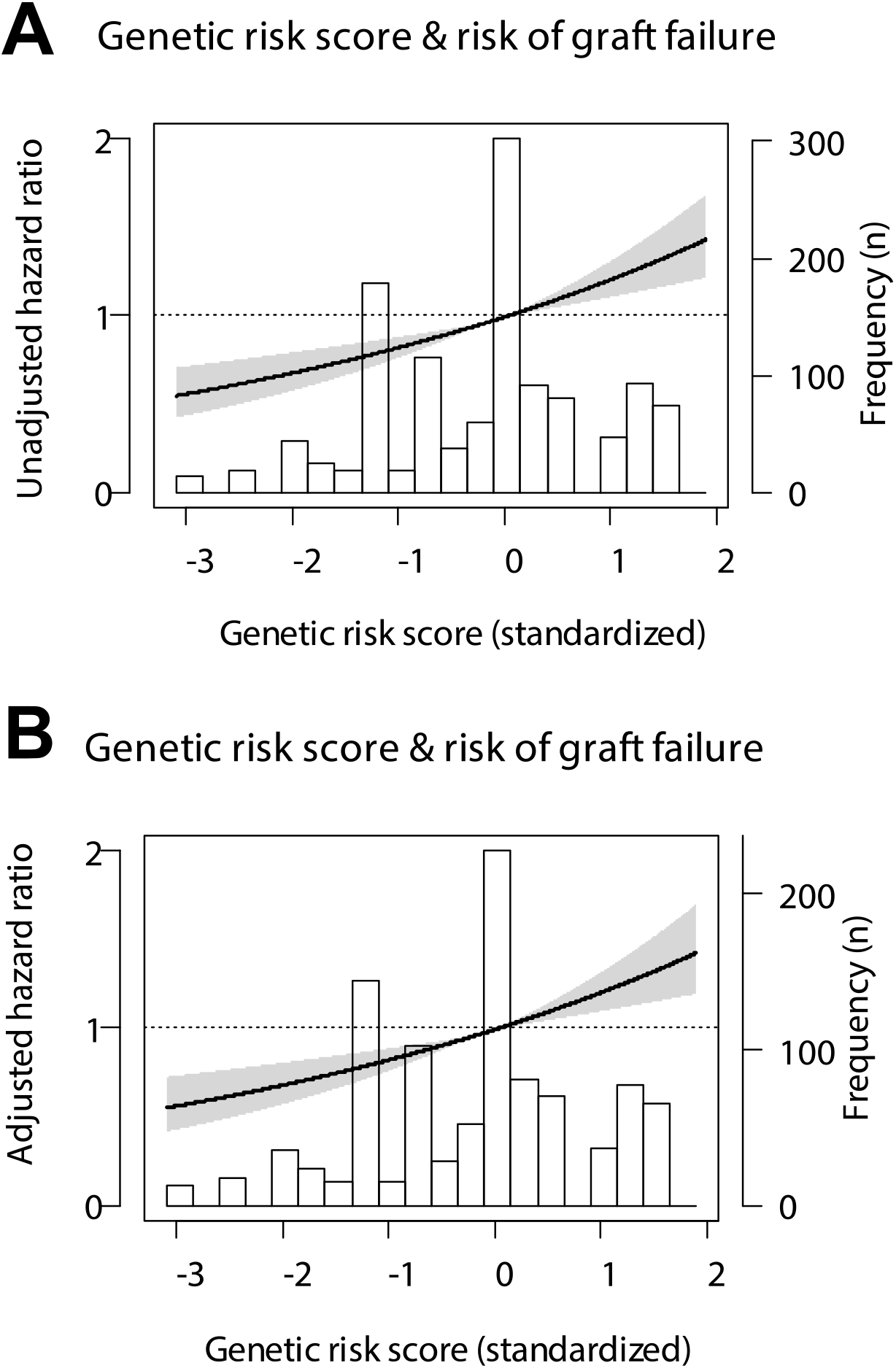
Linear splines of the association of the interleukin-6/interleukin-6 receptor genetic risk score with rejection. Data were fit by a Cox proportional hazard model for all splines (A) unadjusted or (B) adjusted for the recipient age, recipient sex, total number of HLA-mismatches and the occurrence of delayed graft function (DGF). The hazard ratio is represented by the black line, the 95% confidence interval by the gray area.

The performance of the IL-6/IL-6R genetic risk score for the prediction of rejection was also assessed (Table 6). The genetic risk score alone had a Harrell’s C of 0.57 (95% CI: 0.54 – 0.60). Moreover, when added to a model of the *IL6R* SNP in the donor (c-statistic, 0.51; 95%-CI, 0.488 – 0.539), the IL-6/IL-6R genetic risk score significantly increased the Harrell’s C values (c-statistic increase, 0.054; 95%-CI, 0.023 – 0.086; *P* = 0.001). Next, additional variables were included and the discriminative accuracy to predict graft loss of the model improved. The Harrell’s C of the models with the donor characteristics and the transplant variables significantly improved with the addition of the genetic risk score, while only a trend was seen in the model with recipient characteristics. In addition, the IL-6/IL-6R genetic risk score significantly improved the predictive value of the models according to the integrated discrimination improvement index (IDI). Even in the fully adjusted models, the IDI value was >1%, indicating that the IL-6/IL-6R substantially improved risk prediction for rejection markedly beyond currently used clinical markers.

**Table 6.**
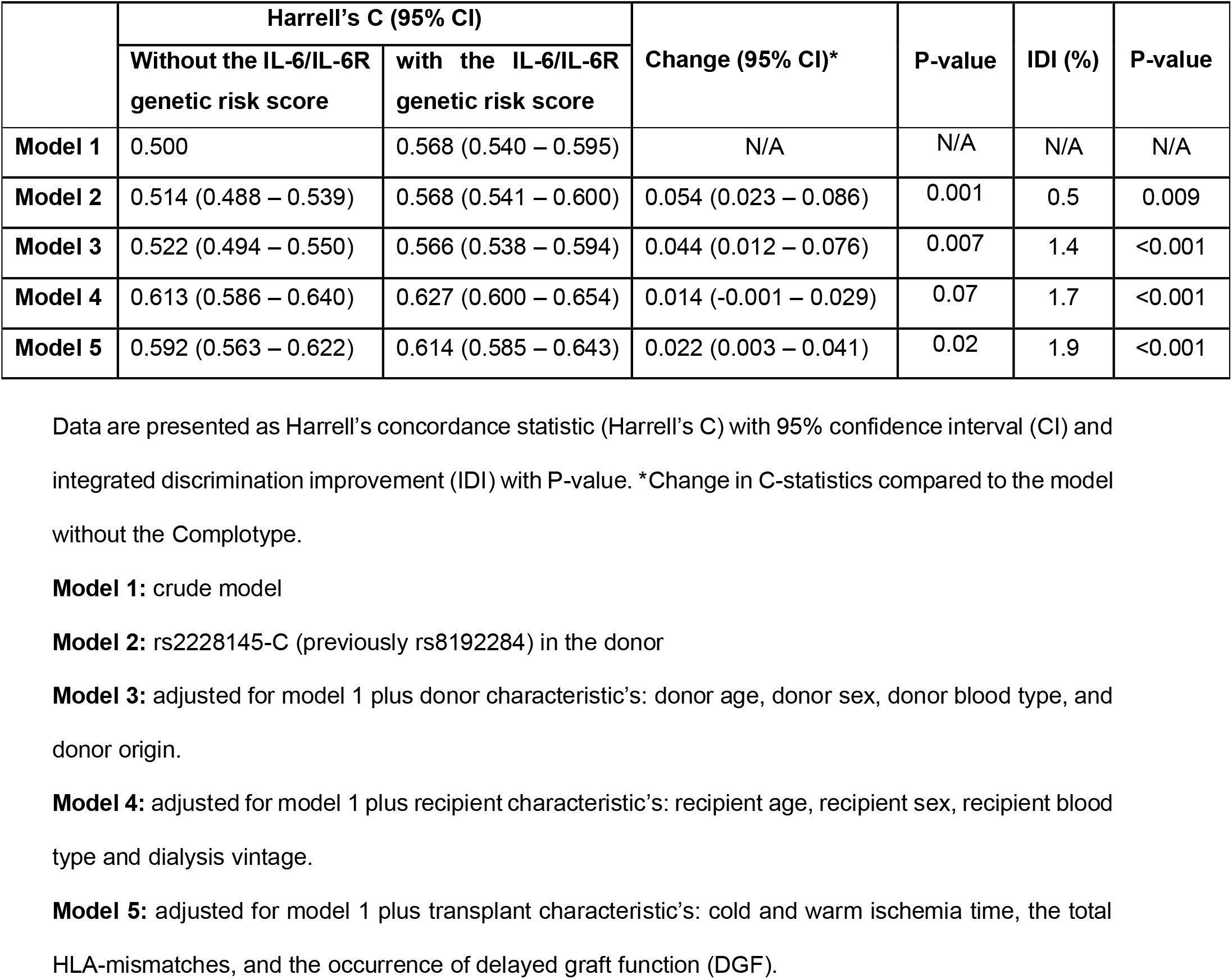
Additive value of the interleukin-6/interleukin-6 receptor genetic risk score in with rejection following kidney transplantation.

## Discussion

Rejection after kidney transplantation remains an important cause of allograft failure that greatly impacts morbidity.^18–20^ Use of novel therapies to reduce allosensitization is vital, but new drug development for kidney transplantation is limited,^21^ necessitating the repurposing of existing anti-inflammatory drugs approved for other indications. Among these are IL-6-blocking therapies, which have already been approved for the treatment of auto-immune diseases.^22^ Human genetics offer an opportunity for target validation. Moreover, drug targets informed by human genetic evidence are more than twice as likely to lead to approved therapeutics.^23,24^ The main finding of our study is a significant association between a common functional *IL6* and *IL6R* polymorphisms in the donor and the risk of biopsy-proven rejection following kidney transplantation. Extending these findings, a genetic risk score based on both SNPs in the donor and recipient was shown to be a major determinant of biopsy-proven rejection. Moreover, this IL-6/IL-6R genetic risk score significantly improved risk prediction for rejection beyond currently used clinical risk factors. In conclusion, our study provides genetic evidence for the potential efficacy of targeting the IL-6 pathway in renal transplantation and encourages the study of IL-6 receptor inhibitors in kidney donors in randomized controlled trials as a donor pretreatment strategy.

To our knowledge, our study is the first to demonstrate that *IL6R* polymorphisms in the donor impact the risk of rejection after kidney transplantation. To sum up, we found that for each copy of the C-allele in the donor the relative risk of biopsy-proven acute rejection decreased by 27% (90%-CI; 14– 38%). In accordance with our results, the same allele (rs2228145-C) has been associated with decreased risks of rheumatoid arthritis and coronary heart disease.^25,26^ A recent study by Bovijn and colleagues found that this *IL6R* variant was also associated with a lower risk of SARS-CoV-2 infection, as well as a lower risk of hospitalization for COVID-19.^27^ Studies on the functional consequences of this variant have helped to understand the molecular mechanisms through which this *IL6R* genotype protects against a wide spectrum of diseases.^28–30^ This non-synonymous polymorphism accounts for over 50% of the total variance in sIL-6R levels and each copy of the C-allele increases plasma levels of sIL-6R. Although these effects of this *IL6R* variant on sIL-6R levels may appear contradictory, further investigation revealed that the C-allele simultaneously reduces membrane-bound IL-6R on monocytes and CD4^+^ T cells (up to 28% reduction per C-allele).^30^ Importantly, reduced surface expression of IL-6R on leukocytes resulted in diminished IL-6 receptiveness, as observed by a reduction in phosphorylation of the key transcription factors STAT1 and STAT3 following stimulation with IL-6.^30^ Additional *in vivo* evidence of the anti-inflammatory effects of this *IL6R* variant has been demonstrated by the association with lower levels of C-reactive protein, fibrinogen, IL-8, and TNF-α in various studies.^26–28,31^ Our analysis convincingly shows that the *IL6R* variant is associated with a lower risk of renal allograft rejection. We postulate that this decrease in rejection rate is due to the amplification of sIL-6R combined with circulating, endogenous, soluble gp130 acting as a buffer to neutralize IL-6, thereby suppressing inflammation as well as decreasing allosensitization, invariably diminishing the risk of allograft rejection.

The impact of the *IL6* ∼174G/C polymorphism (rs1800795) on the risk of rejection among transplantation patients has previously been investigated by several studies.^12,32–36^ A recent metanalysis of seven studies including 369 cases and 679 controls concluded that the recipient *IL6* genotype was not significantly associated with rejection, whereas a trend was seen for donor *IL6* genotypes.^13^ In our transplant cohort of 1,271 patients we did not find such an association either. Yet, when we performed a subgroup analysis for the sex of the donor, we observed a significant association between the *IL6* SNP and rejection in male donors. This sex-related difference might also explain the conflicting results by previous studies. Similar to our observations, others have reported a clear sex dimorphism in the association of this *IL6* polymorphism with the vulnerability to illnesses.^37–39^ Overall, the associations with this *IL6* variant were stronger and predominantly found in men. Our study, therefore, highlights that sex should be taken into account in transplantation as well as for cytokine-targeted therapies. Furthermore, in conformity with our results, the C-allele of this *IL6* SNP has also been associated with increased risks of rheumatoid arthritis and cardiovascular disease in recent meta-analyses.^40–42^ For rs1800795, the G-allele was initially said to increase IL-6 levels, however, recent work revealed that the C-allele of this SNP leads to higher IL-6 expression in fibroblasts but not in leukocytes.^43^ Brull *et al*. found different kinetic profiles for IL-6 increase after surgery based on *IL6* genotypes, which could explain previous conflicting results.^44^ Nevertheless, the overall increase in IL-6 was more profound in CC homozygotes. Further *in vivo* evidence of the pro-inflammatory effects of the C-allele is demonstrated by the association with higher serum levels of IL-6, C-reactive protein, and fibrinogen in multiple studies.^45–47^ Altogether, our study adds to a growing body of evidence that connects local production of IL-6 to the allosensitization of recipients to renal allografts.^48,49^

Tocilizumab, a humanized monoclonal antibody targeting the IL-6R, has been assessed for the treatment of acute rejection, chronic ABMR, and transplant glomerulopathy following renal transplantation.^50–52^ Initially, a phase I/II trial was performed in 10 patients prior to kidney transplantation that were unresponsive to desensitization with intravenous Ig and rituximab.^50^ Tocilizumab, combined with intravenous Ig, led to a significant reduction in DSA levels and appeared safe. Five patients were transplanted and six-month protocol biopsies showed no acute rejection or transplant glomerulopathy. Next, an open-label single-center trial was undertaken in 36 patients with chronic ABMR that were non-responsive to intravenous Ig and rituximab.^51^ Patient and renal allograft survival was 91% and 80% at 6 years, respectively, and this was found to be superior to historical controls. Furthermore, stabilization of renal function was seen after 2 years. Finally, tocilizumab was investigated for the treatment of acute rejection in an observational study of 7 kidney transplant recipients.^52^ Renal function stabilized or improved in all patients, but one patient had a potential hypersensitivity reaction, and another patient developed cytomegalovirus esophagitis. A multicenter randomized control trial is currently underway.^53^ Considering IL-6 inhibitors are already being tested in clinical trials, what, then, is the role for genetic studies here? The results from our study provide important considerations for the design of these clinical trials by identifying the following key issues for targeting IL-6 in renal transplantation: (i) Site of action; (ii) Timing of treatment; (iii) Sex differences; and, (iv) Patient selection.

We found that genetic polymorphisms of the IL-6 signaling pathway in donors, but not recipients, were associated with the risk of biopsy-proven rejection after kidney transplantation. These results indicate that not circulating IL-6 in the recipient but local IL-6R expression and IL-6 production by the donor kidney are key drivers of allosensitization to the kidney transplant. This potentially suggests that therapies aimed at blocking IL-6/IL-6R interactions should focus on the donor kidney as the site of action. Neutralizing IL-6R nanobodies (i.e., ALX-0061) could therefore be more effective than larger anti–IL-6R (i.e., tocilizumab) or anti–IL-6 antibodies (i.e., clazakizumab), due to better tissue penetration of the donor kidney.^4^ Alternatively, the use of small interfering RNA (siRNA) could be a promising approach.^54,55^ The found association we found between donor genetics and transplant outcomes also indicates that donor pre-treatment might be the optimal timing of treatment. Interventions within the donor may protect against a proinflammatory reaction after transplantation, and therefore, donor pre-treatment could be a promising strategy to increase graft function and survival.^56^ Conversely, donor pre-treatment is not possible until it is known whether other donor organs (i.e., the liver, heart, and lungs) also benefit from targeting IL-6. Instead, treatment could be given during the preservation phase (i.e., cold storage or machine perfusion), thereby targeting the donor kidney specifically. Finally, in this study, we constructed an IL-6/IL-6R genetic risk score based on two SNPs in the donor and recipient. From a prognostic perspective, this genetic risk score could be used for the prediction of patients at high risk of allograft rejection. Furthermore, the genetic risk score may be used to identify renal transplant pairs that could benefit from anti-IL-6 treatment.

Several limitations of our study warrant consideration. First, the associations found in this study are expected to be causal. However, since our study is prospective, but observational in nature, it cannot be proven by our results. Second, we only performed an analysis of individual functional SNPs and not for *IL6* and *IL6R* haplotypes. Third, measurements of IL-6 and sIL-6R were not performed in our cohort due to the lack of serum samples, and genotypes could therefore not be correlated to systemic levels. Finally, we could not investigate whether the association between the *IL6* and *IL6R* variants differed for TCR or ABMR, due to the lack of a standardized assay over the years for the determination of DSA. On the other hand, major strengths of our study are the large sample size, robust statistical analysis (incl. subgroup analysis), and the hard and clinically relevant endpoint, namely biopsy-proven rejection.

In conclusion, we found that *IL6* and *IL6R* variants in the donor associate with the risk of developing biopsy-proven rejection after renal transplantation. These findings imply potential efficacy of targeting IL-6 signalling in renal transplantation. Ongoing, randomized controlled trials with IL-6 or IL-6R inhibitors are needed to identify the best settings, including the timing of intervention and patient selection, in which these agents might be effective.

## Methods

### Subjects

We enrolled patients who underwent single kidney transplantation at the University Medical Center Groningen in the Netherlands between March 1993 until February 2008. From the 1430 renal transplantations, 1271 recipient and donor pairs were included in the cohort as previously described.^57– 59^ Subjects were excluded due to technical complications during surgery, lack of DNA, re-transplantation or loss of follow-up. This study is in accordance with the declaration of Helsinki and all patients provided written informed consent. The medical ethics committee of the University Medical Center Groningen approved the study under file n° METc 2014/077.

### DNA isolation and genotyping

Peripheral blood mononuclear cells were isolated from blood or splenocytes collected from the donors and recipients. DNA was extracted with a commercial kit as instructed by the manufacturer and stored at −80°C. Genotyping of the SNPs was determined via the Illumina VeraCode GoldenGate Assay kit (Illumina, San Diego, CA, USA), according to the manufacturer’s instructions. The promoter of IL6 contains several SNPs, of which the rs1800795-174 G>C is the most widely studied for its influence on acute rejection of renal allograft.^13^ In addition, we chose the *IL6R* rs2228145 A>C (formerly rs8192284) SNP, which has previously shown to impair IL-6R signaling and influence the risk of diverse inflammatory diseases.^30^ Genotype clustering and calling were performed using BeadStudio Software (Illumina). The overall genotype success rate was 99.5% and 6 samples with a high missing call rate were excluded from subsequent analyses.

### Study end-points

The primary end-point in this study was biopsy-proven rejection (all biopsies were re-evaluated according to the Banff 2007 classification) after transplantation.

### Genetic risk score

We created a genetic IL-6 and IL-6R risk score that assigns points for the presence of a risk-decreasing or a risk-increasing allele in the donor and recipient. However, to take into account the strength of the association of the SNPs with rejection, the point for the presence of an *IL6* or *IL6R* SNP is multiplied by the regression coefficient (= logarithm of the hazard ratio) creating a weighted risk score. A regression coefficient is negative when an SNP is protective and the regression coefficient is positive when an SNP is hazardous. The total sum of the *IL6* or *IL6R* SNPs in both the donor and recipient creates the genetic risk score. Next, we determined whether the genetic risk score improved the prediction of rejection compared to only the *IL6R* SNP in the donor.

### Statistical analysis

Statistical analyses were performed using SPSS version 25. Data are displayed as median [IQR] for non-parametric variables; mean ± standard deviation for parametric variables and the total number of patients with percentage [n (%)] for nominal data. Differences between groups were examined with the Mann-Whitney-U test or the Student t-test for not-normally and normally distributed variables, respectively, and categorical variables with the χ2 test. Log-rank tests were performed between groups to assess the difference in the incidence of biopsy-proven rejection. Univariable analysis was performed to determine the association of genetic, donor, recipient, and transplant characteristics with rejection. The factors identified in these analyses were thereafter tested in a multivariable Cox regression. Additionally, multivariable cox regression with a stepwise forward selection was performed. Harrell’s C statistic was used to assess the predictive value of the SNPs when added to the reference model. The additional value of the genetic risk score was assessed by the integrated discrimination improvement (IDI). The IDI indicates the difference between model-based probabilities for events and non-events for the models with and without the genetic risk score. Tests were 2-tailed and regarded as statistically significant when *P*<0.05.

## Data Availability

For original data, please contact

## Author contributions statement

Study design by JD and MS. Acquisition of data by JD and FP. Analysis and interpretation of data by FP, MG and JD. Drafting of manuscript by FP, MG and SE. All authors were involved in editing the final manuscript. All authors read and approved the final manuscript.

## Acknowledgement

The authors thank the members of the REGaTTA cohort (REnal GeneTics TrAnsplantation; University Medical Center Groningen, University of Groningen, Groningen, the Netherlands): S. J. L. Bakker, J. van den Born, M. H. de Borst, H. van Goor, J. L. Hillebrands, B. G. Hepkema, G. J. Navis and H. Snieder. Furthermore, we thank Camilo Sotomayor Campos for the preparation of Figure 5.

## Abbreviations

AUC: Area under the curve
ABMR: Antibody mediated rejection
CIT: Cold ischemia time
DBD: Donation after circulatory death
DCD: Donation after brain death
DGF: Delayed graft function
ESKD: End-stage kidney disease
Gp130: Glycoprotein 130 (gp130)
HLA: Human leukocyte antigen
HR: Hazard ratio
IDI: Integrated discrimination improvement
IL 6: Interleukin 6
*IL6*: Interleukin 6 gene
IL-6R: Interleukin 6 receptor
*IL6R*: Interleukin 6 receptor gene
JAK: Janus kinase (JAK)
mIL-6R: Membrane-bound interleukin 6 receptore
PRA: Panel-reactive antibody
STAT: Signal transducer and activator of transcription
sIL-6R: Soluble Interleukin 6 receptor
SNP: Single-nucleotide polymorphism
WIT: Warm ischemia time

